# Evaluation of real-life use of Point-Of-Care Rapid Antigen TEsting for SARS-CoV-2 in schools (EPOCRATES)

**DOI:** 10.1101/2021.10.13.21264960

**Authors:** Ana C. Blanchard, Marc Desforges, Annie-Claude Labbé, Cat Tuong Nguyen, Yves Petit, Dominic Besner, Kate Zinszer, Olivier Séguin, Zineb Laghdir, Kelsey Adams, Marie-Ève Benoit, Geneviève Leduc, Jean Longtin, Ioannis Ragoussis, David L. Buckeridge, Caroline Quach

## Abstract

**Abstract:** *Background:* We evaluated the use of rapid antigen detection tests (RADT) for the diagnosis of severe acute respiratory syndrome coronavirus 2 (SARS-CoV-2) infection in school settings to determine RADT’s performance compared to PCR.

*Methods:* In this real-world, prospective observational cohort study, high-school students and staff were recruited from two high-schools in Montreal (Canada) and followed from January 25^th^ to June 10^th^, 2021. Twenty-five percent of asymptomatic participants were tested weekly by RADT (nasal) and PCR (gargle). Class contacts of cases were tested. Symptomatic participants were tested by RADT (nasal) and PCR (nasal and gargle). The number of cases and outbreaks were compared to other high schools in the same area.

*Results:* Overall, 2,099 students and 286 school staff members consented to participate. The overall RADT’s specificity varied from 99.8 to 100%, with a lower sensitivity, varying from 28.6% in asymptomatic to 83.3% in symptomatic participants. Secondary cases were identified in 10 of 35 classes. Returning students to school after a 7-day quarantine, with a negative PCR on D6-7 after exposure, did not lead to subsequent outbreaks. Of cases for whom the source was known, 37 of 57 (72.5%) were secondary to household transmission, 13 (25%) to intra-school transmission and one to community contacts between students in the same school.

*Conclusion:* RADT did not perform well as a screening tool in asymptomatic individuals. Reinforcing policies for symptom screening when entering schools and testing symptomatic individuals with RADT on the spot may avoid subsequent significant exposures in class.

*Table of Contents Summary:* Rapid antigen tests were compared to standard PCR to diagnose SARS-CoV-2 infections in high-school students. They performed better in symptomatic individuals.

*What’s Known on This Subject:* Rapid antigen detection tests (RADT) are often used to diagnose respiratory pathogens at the point-of-care. Their performance characteristics vary, but they usually have high specificity and moderate sensitivity compared with PCR.

*What This Study Adds:* RADT sensitivity ranged from 28.6% in asymptomatic individuals to 83.3% in symptomatic individuals. Return to school after 7 days of quarantine was safe in exposed students. Secondary cases were identified in 28% of classes with an index case.

## Background

Timely diagnosis of infection enables outbreak control through rapid isolation of index cases and subsequent contact tracing (1, 2). Diagnosis of severe acute respiratory syndrome coronavirus 2 (SARS-CoV-2) infection is predominantly based on polymerase chain reaction (PCR), which has a turnaround time (TAT) of 24-48 hours. Rapid antigen detection tests (RADT) are inexpensive and can be used at the point-of-care. They usually have high specificity and moderate sensitivity compared with PCR (3-6). Given their rapid TAT, RADT allow for efficient triage and management of exposed individuals (7). The potential use of RADT is especially relevant in schools, where COVID-19 outbreaks can interrupt in-person teaching and negatively impact learning (8-11).

RADT perform best in the early stages of infection, when viral load is generally high (12-15). Reported RADT sensitivity ranges from 28.9% to 98.3%, with improved RADT sensitivity in samples with high viral loads and in symptomatic individuals (16, 17). The usual limits of detection (LOD) for PCR is 600-1000 viral RNA copies/ml, whereas RADT usually have LOD 2-3 logs higher (10^5^ to 10^6^) (18). Many studies have indicated the importance of high viral load dynamics with infectiousness (19, 20). For each unit increase in Ct value, the odds of recovering infectious virus decreased by 0.67, being under 10% when Ct-values were >lJ35. Ct values of 17 to 32 corresponded to 10^5^ and 10^1^ SARS-CoV-2 RNA copies/µL, respectively (21).

We aimed: 1) to determine the performance characteristics of RADT for SARS-CoV-2 compared to PCR in high-school students and staff and 2) to determine if serial testing of COVID-19 contacts would allow for safe faster return to school.

## Methods

### Participating population

The study was conducted in two high schools of Montreal. Pensionnat du Saint-Nom-de-Marie (PSNM) is a private school, with most students from native-born affluent families. École secondaire Calixa-Lavallée (ESCL) is a public school where students are predominantly from first-generation immigrant communities. Both schools followed the Ministry of Education recommendations, by forming “classroom bubbles”. Masks were mandatory as of October 8^th^, 2020. Students were ∼30/class and seated three feet apart. School staff were invited to participate. Vaccination began April 9^th^, 2021 in adults and May 25^th^, 2021 for children ≥12 years.

### Study design and interventions

This was a real-world, prospective observational cohort study comparing RADT to PCR, from January 25^th^ to June 10^th^, 2021.

The lateral flow immunoassay [PanBio™ COVID-19 Ag test (Abbott Laboratories, Illinois, USA)], authorised by Health Canada (22) was used. Nasal swabs were self-collected under the supervision of a research assistant, to avoid sampling bias, who then performed RADT on site. Spring water gargle specimens were collected for PCR testing (23). Laboratory-developed PCR was performed at CHU Sainte-Justine, with a LOD of 400 copies/mL (24). Extraction and purification of genetic material was done with Roche’s MagNA Pure 96 system. The laboratory testing protocol and the water gargle validation have been described elsewhere (25-28).

Decisions about management of cases and contacts were made by two members of the research team (AB, CQ), in collaboration with local public health (CT, OS). The school principals (YP, DB) were actively involved in the study deployment.

#### 1) Testing protocol in the absence of a known exposure

a. Asymptomatic individuals: Nasal swabs and gargle specimens were collected weekly for RADT (nasal) and PCR (gargle) on a random sample of 25% of participants, stratified by class.
b. Symptomatic individuals: Gargle specimens for PCR and a nasal swab for RADT and PCR were performed on site. Results from RADT and PCR were reported to public health; an individual was considered infected if the PCR result was positive. If symptoms occurred in school, the research team proceeded with testing. If symptoms developed at home, participants could get tested at school in a private room.

#### 2) Management of exposed contacts of a positive individual in a class

Contacts of a confirmed positive individual were isolated at home. Students were allocated to a 7- or 14-day quarantine, staffs were allocated to a 7-day quarantine, with tests (nasal RADT and gargle PCR) three days after last contact with the known positive case, and up to two days before the end of quarantine. RADT was performed on day (D)14, D21 and D28, if the initial PCR was negative. If symptoms developed, both the RADT and PCR were performed. Students who did not consent to the study were quarantined for 14 days. Students and staff with significant off-campus exposures were offered on site testing.

### Outcomes

The primary outcome was to assess the performance characteristics of RADT in: a) asymptomatic participants randomly screened; b) asymptomatic close contacts of a confirmed positive case; c) symptomatic participants.

Secondary outcomes included: a) number of positive students by RADT in groups exposed to a confirmed positive index case, allocated to early (on D8) versus standard (on D15) return to school and b) number of case clusters in schools. This was compared to clusters in other high schools in Montréal during the same time frame, using public health data.

### Statistical analysis

Descriptive statistics were used for the performance of the RADT. To determine the precision with which we could estimate our primary outcome, we implemented an agent-based model (ABM) (29) (Supplementary Appendix A). Based on this simulation, we expected that the number of infections and tests would be sufficient in one school but added a second school to support generalizability of the findings and explore secondary objectives.

### Ethical considerations

This project was approved by the CHU Ste-Justine Research Ethics Board (#MP-21-2021-3271). Informed parental consent or assent were required for all students. Parents who preferred to keep their children home for 14 days in case of a contact could do so. Tests results were communicated to parents and students by the school. This study was funded by the Québec Ministry of Health and Social Services.

## Results

During the study period, 2,099 students and 286 school staff members consented to participate. The participation rate for students was 78.5% and 63.5% (Figure 1) and 94.4% and 89.5% for staff.

**Figure 1:**
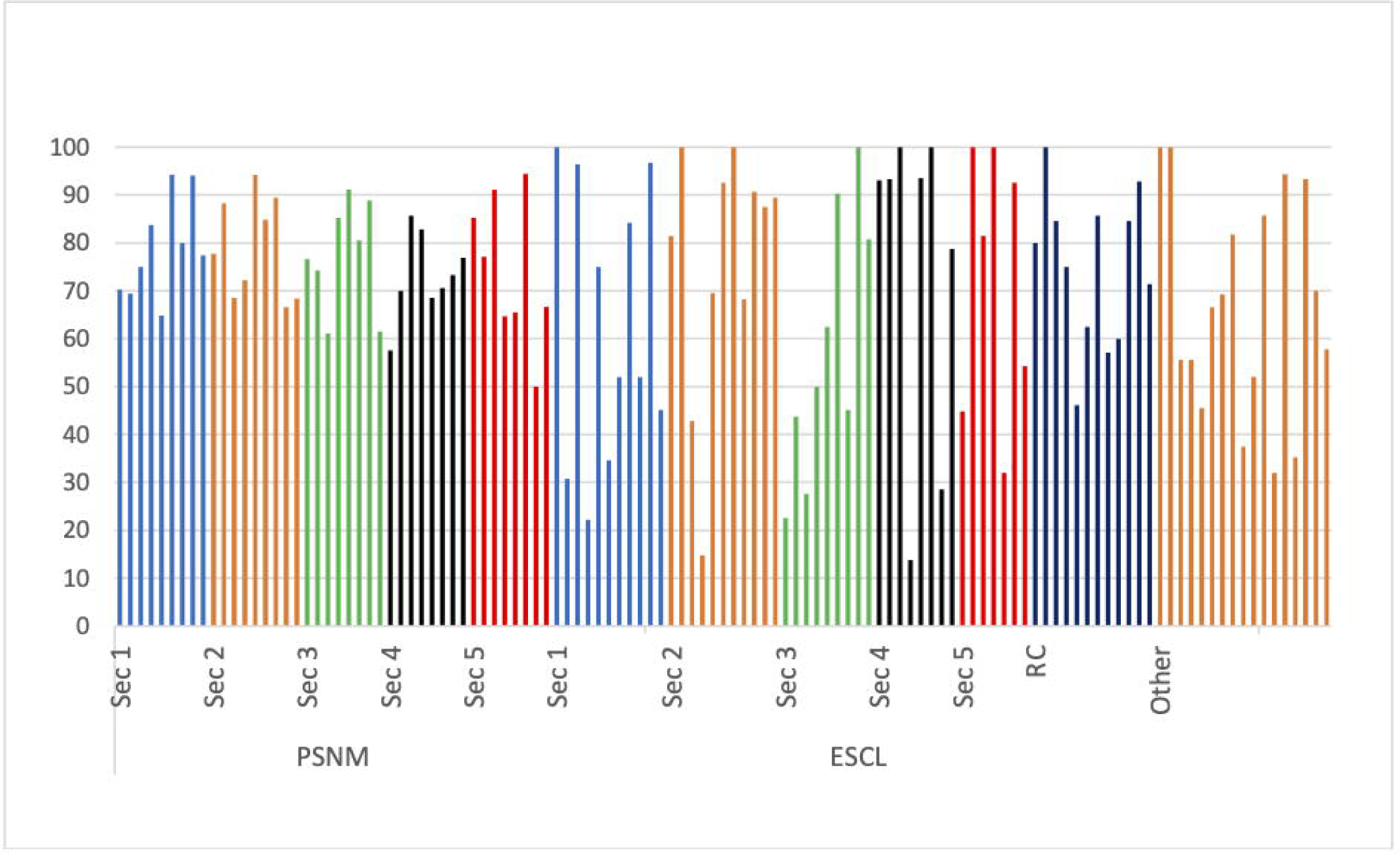
Proportion of participating students per class and level. Sec: Secondary; RC: Reception class; PSNM: Pensionnat du Saint-Norn-de-Marie; ESCL : École Secondaire Calixa-Lavallée.

### RADT results and PCR validation (from gargle specimens only)

#### 1) Asymptomatic students and staff

Of 5,583 RADT done on asymptomatic students (Table 1), seven had an invalid PCR result on the gargle sample, seven were equivocal and three were weak positive, of which one was negative when repeated the next day (and was excluded). Two students with equivocal or weak positive PCR results had a positive PCR result in the previous 90 days. The prevalence in asymptomatic participants was 0.30% (95% CI 0.18-0.49). Therefore, the sensitivity of RADT in that population was 41.2% (95% CI 21.6-64.0), with a specificity of 100.0%.

**Table 1.** Performance of RADT in the different participant groups.

| RADT<br>(nasal) | RESULTS |  |  |  | CLINICAL PERFORMANCE OF RADT |  |  |
| --- | --- | --- | --- | --- | --- | --- | --- |
|  | PCR (gargle) |  |  |  | Sensitivity |  | Specificity |
|  | POS | NEG | EQ/Weak POS | INV | Excluding EQ/Weak POS | Including EQ / Weak POS |  |
| *Asymptomatic students\** |  |  |  |  |  |  |  |
| POS | 7 | 1 | 0 | 0 | 41.2% | 26.9% | 100%† |
| NEG | 10 | 5549 | 9 | 7 | (95% CI 21.6-64.0) | (95% CI= 13.7-46.1) | (95% CI 99.9-100) |
| INV | 0 | 0 | 0 | 0 | (n=17) | (n=26) |  |
| *Asymptomatic students considered exposed contacts of positive index cases‡* |  |  |  |  |  |  |  |
| POS | 4 | 6 | 0 | 0 | 28.6% | 26.7% | 99.6% |
| NEG | 10 | 1470 | 1 | 0 | (95% CI 8.4-58.1) | (95% CI 7.8-55.1) | (95% CI 99.1-99.9) |
| INV | 0 | 0 | 0 | 0 | (n=14) | (n=15) |  |
| *Symptomatic students§* |  |  |  |  |  |  |  |
| POS | 10 | 0 | 0 | 0 | 83.3% |  | 100.0% |
| NEG | 2 | 223 | 0 | 0 | (95% CI 51.6-97.9) | N/A | (95% CI 98.4-100.0) |
| INV | 0 | 0 | 0 | 0 |  |  |  |
| *Asymptomatic staff members* |  |  |  |  |  |  |  |
| POS | 0 | 1 | 0 | 0 |  |  | 99.8% |
| NEG | 0 | 775 | 0 | 0 | N/A | N/A | (95% CI 99.3-100.0) |
| INV | 0 | 0 | 0 | 0 |  |  |  |
| *Asymptomatic staff members considered exposed contacts of positive index cases* |  |  |  |  |  |  |  |
| POS | 0 | 0 | 0 | 0 |  |  | 100.0% |
| NEG | 0 | 61 | 0 | 0 | N/A | N/A | (95% CI 94.1-100.0) |
| INV | 0 | 0 | 0 | 0 |  |  |  |
| *Symptomatic staff members* |  |  |  |  |  |  |  |
| POS | 1 | 0 | 0 | 0 | 50.0% |  | 100.0% |
| NEG | 1 | 62 | 0 | 0 | (95% CI 1.3-98.7) | N/A | (95% CI 94.3-100.0) |
| INV | 0 | 0 | 0 | 0 |  |  |  |

RADT: rapid antigen detection test, PCR: polymerase chain reaction, POS: positive, NEG: negative, INV: invalid, EQ: equivocal, CI: confidence interval; N/A: non-applicable
\* Prevalence of SARS CoV-2 infection, based on PCR results (including equivocal and weakly positive results): 0.30% (95% CI 0.18-0.49)
†The specificity of RADT in asymptomatic students was 99.98% when adjusted to two decimal places
‡ Prevalence of SARS CoV-2 infection, based on PCR results (including equivocal and weakly positive results): 0.7% (95% CI 0.5-1.6)
§ Prevalence of SARS CoV-2 infection, based on PCR results (including equivocal and weakly positive results): 5.1% (95% CI 2.65-8.71)

Of 784 asymptomatic RADT screening tests done on asymptomatic randomly screened staff members, two gave invalid PCR results and six were lost. Only one RADT was positive, but the PCR was negative on both the gargle and nasal specimens, giving a specificity of 99.8% (95% CI 99.3-100.0) (Table 1).

#### 2) Asymptomatic exposed contacts in a classroom

A total of 1491 RADT tests and PCR were done on asymptomatic students exposed to a positive classmate index case at D3 and 2 days before returning to class. After excluding one equivocal PCR result, SARS-CoV-2 prevalence in this exposed group was 0.7% (95% CI 0.5-1.6). The sensitivity of RADT was 28.6% (95% CI 8.4-58.1) with a specificity of 99.6% (95% CI 99.1-99.9) (Table 1). Of 627 RADT done for asymptomatic exposed contacts on D14, D21 and D28, only one was positive (also positive by PCR when tested on D12 – see below). A total of 61 RADT and PCR were done for staff members on D3 and D7 after a contact with a positive index case in school (Table 1). All were negative.

#### 3) Symptomatic students and staff

Overall, 235 students developed symptoms and were tested on site for SARS-CoV-2. As shown in Table 1, 10 had a positive RADT and 12 had a positive PCR [prevalence=5.1% (95% CI 2.7-8.7)]. The sensitivity of RADT in that population was 83.3% (95% CI 51.6-97.9) with a specificity of 100.0% (95% CI 98.4-100.0). Sixty-four staff members were tested on site for symptoms compatible with COVID-19. Only one had a positive RADT and PCR. One positive case was identified by PCR after a negative RADT (sensitivity of 50% (95% CI 1,3-=98,7) and specificity of 100%.

### Outbreaks and comparison with other schools in the region

We identified 76 PCR (gargle or nasal) positive cases, including three cases in staff. Of the 35 classes included in the study, 20 returned on D8 after contact, if the gargle PCR was negative on D6 or D7.

Secondary cases were identified in 10 classes. The number of secondary cases in each class were one (n=8 classes), three (n=1 class) and four (n=1 class). Four secondary cases had a positive RADT, including three asymptomatic students and one symptomatic student who tested positive by RADT and PCR on D12, with symptoms starting on D9 after last contact with the positive classmate – a community exposure was also suspected. No tertiary case occurred. Outbreaks were limited to the classroom bubble and to school friends seen outside of school. When the source was known, 37/57 cases (72.5%) were secondary to household transmission, 13 (25%) to intra-school transmission and one to community contacts between students in the same school.

During the same period, outbreaks declared in other Montreal schools had a lower proportion of asymptomatic cases (31.8%) compared to ESCL (55.6%) and PSNM (85.7%) (Supplementary Appendix B).

## Discussion

RADT were purchased worldwide as a tool to prevent outbreaks. However, their use is limited by the paucity of evidence regarding their performance in children. In this study, we prospectively compared the performance of a RADT to PCR for the purpose of limiting transmission of SARS-CoV-2 infection in schools. In a context of lower SARS-CoV-2 prevalence in school than in the community (30), we observed only seven false positive RADT during the 5-month study (all in asymptomatic individuals) and the specificity of the RADT remained overall excellent. However, the sensitivity was much lower, varying between 28.6% in asymptomatic to 83.3% in symptomatic students.

A recent large observational study described the use of RADT in asymptomatic individuals as beneficial, reporting a sensitivity of 64.4% (95% CI 58.3-70.2) (31). However, this could be overestimated as not all asymptomatic individuals had a confirmatory PCR. In our study, only a few positive cases were detected by RADT (overall 7/6358, 0.11%) in asymptomatic individuals who were randomly tested. Ten additional cases were detected by PCR from gargle specimens. Two full-time research assistants were in each school, in addition to local school staff who were supporting the study rollout. This level of required resources goes against the use of RADT for random screening of asymptomatic individuals, given low sensitivity in that setting.

RADT detected SARS-CoV-2 positive symptomatic cases in 15 minutes, allowing for prompt isolation, contact tracing and testing. The overall sensitivity of RADT in symptomatic individuals was 78.6% (95% CI 49.2-95.3). This finding is in agreement with other published studies (14, 15, 32-34). Sood et al. recently described that the positive concordance of RADT was higher among symptomatic children (64.4%) compared to asymptomatic children (51.1%) presenting at a walk-in testing site (33). L’Huillier et al. described a sensitivity of 73.0% in symptomatic vs. 43.3% in asymptomatic children (34). The authors described the peak of sensitivity on D2 post symptoms onset, with a subsequent decrease to 56% by D5. In our study, 225 of 235 symptomatic children had recorded their symptoms onset, with a median time of one day (range: 0-33 days). Overall, 46.7% (n=105/225) were tested with RADT and PCR on the day of symptoms onset. Our reported RADT sensitivity may have been higher had students been tested on subsequent days. However, the usefulness of RADT is to control outbreaks, therefore delaying testing to enhance sensitivity would be counterproductive. This trade-off may not apply to the Delta variant, for which the kinetic of infection may differ (35, 36).

RADT identified 28.6% of positive asymptomatic exposed school contacts, which was similar to that recently described by Torres et al. for non-household significant contacts (sensitivity: 35.7%) (37). Although this percentage is low, the rapid diagnosis of SARS-CoV-2 infection in exposed individuals allowed local public health to quickly manage these students’ household contacts who, at the time, had to isolate until the result of the D3 testing. With changes in quarantine recommendations for vaccinated family members, the benefit of RADT in this group may be smaller. Most positive cases in students were due to household SARS-CoV-2 transmission. Students were often sent to school despite having a known positive contact. Active screening of symptoms and history of significant exposures should be reinforced to prevent school outbreaks. Thirteen of 51 cases were acquired from school, with 15 cases in the same class bubble (in five classes overall). Therefore, the asymptomatic nature of this infection makes screening for school contacts essential. Our results demonstrate that using a more sensitive method, such as PCR, may be more reliable for that purpose.

This study had several limitations. We did not collect data regarding adherence to public health measures, nor systematically documented exposures occurring outside of school. However, for the most part, we were able to identify when significant household transmission occurred and relied on the transparency of participants. We cannot infer whether PCR positive individuals were contagious. The study was performed before the advent of the Delta variant in our region. Because RADT detects the N protein, we expect that its sensitivity and specificity would not be affected negatively, as viral loads of Delta variant infections are reported to be higher (35). Recently published data indicates that the performance of the PanBio™ RADT is similar for detection of the Delta variant than for other variants (38). Despite vaccination, transmission of SARS-CoV-2 is occurring in schools, therefore the findings of this study related to the use of RADT to prevent outbreaks are valid and relevant. Finally, the sensitivity of RADT in symptomatic individuals was based on a relatively small number of people with PCR-confirmed SARS-CoV-2 infection.

This is the largest study to date assessing the use of RADT in schools. The strengths of this study included its prospective design and the real-world use of RADT versus PCR. We assigned participants to earlier versus standard return to school with serial RADT, showing that there were no secondary outbreaks with shorter quarantine. Although the study was not powered to rule this out, this aligns with other recently published data (39) and may allow policymakers to consider reducing the duration of quarantine for exposed contacts, provided a PCR is negative on D6 or D7.

In conclusion, our findings contribute to the growing evidence that the use of RADT leads to rapid diagnosis of SARS-CoV-2 infection in symptomatic individuals (40). However, RADT does not perform as well as a screening tool in asymptomatic individuals. In our study, teenagers were able to proceed to self-collection of swabs, while supervised by a research assistant. It may be helpful to reinforce policies for symptom screening when entering schools, where symptomatic individuals could be tested with RADT to avoid significant in-class exposures. A negative RADT could still mean that symptoms are due to SARS-CoV-2, but with a viral load too low to be detected and therefore less likely to transmit at that point. In such instance, a subsequent sample tested by PCR would be useful.

## Supporting information

Sample size calculation

Comparison of outbreaks to other high schools in Montreal using public health data during the same period

Predicted days of isolation

Distribution of non-index cases linked to an outbreaks according to time to symptoms

## Data Availability

All data produced in the present study are available upon reasonable request to the authors

## Abbreviations

ABM: (agent-based model)
Ct: (cycle threshold)
COVID-19: (Coronavirus disease 2019)
D: (day)
ESCL: (École Secondaire Calixa-Lavallée)
LOD: (limit of detection)
PCR: (polymerase chain reaction)
PSNM: (Pensionnat du Saint-Nom-de-Marie)
RADT: (rapid antigen detection test)
SARS-CoV-2: (Severe acute respiratory syndrome coronavirus 2)
TAT: (turnaround time)

## Notes

**Conflict of Interest Disclosures:** The authors declare that they have no known competing financial interests or personal relationships that could have appeared to influence the work reported in this paper.

### Competing Interest Statement

The authors have declared no competing interest.

### Funding Statement

This study was funded by the Quebec Ministry of Health and Social Services, to whom regular reports of the study progress were submitted. The study sponsor did not have a role in study design, in interpretation of data, writing of the report or decision to submit the paper for publication.

### Author Declarations

This project was approved by the CHU Ste-Justine Research Ethics Board (#MP-21-2021-3271). Written invitation letters to participate in the study were sent by schools direction to parents and staff explaining the study objectives, methods and expected impacts. Online informed parental consent, as well as assent, was required for all students. Parents who preferred to keep their children home for 14 days in case of a class contact could do so. Tests results were communicated to parents and students (if ≥14 years) by the school (via email), as they became available.

### Summary of Updates

Updated version of manuscript, including a correction to Table 1.

