## Supplementary material for "Evaluation of real-life use of Point-Of-Care Rapid Antigen TEsting for SARS-CoV-2 in schools (EPOCRATES)": Sample size calculation

***Supplementary Appendix A : Sample size calculation***

As the school populations were fixed in size, we determined the precision with which we would be able to estimate our primary outcome. Given the non-linear nature of the epidemic process and the complexity of the quarantine and testing policies proposed, it was not possible to estimate precision through a direct calculation. We therefore implemented an agent-based model (ABM) to estimate through simulations the number of tests that would be performed, the likely results of the tests, and other outcomes of interest in planning the study (e.g. number of days in school, number of secondary infections). We implemented a variation of a previously described school-based ABM to adapt the characteristics of the school^29^. For simplicity, we did not model household transmission explicitly, although we did allow for infection outside of the school. We also extended the model to include testing and quarantine, and we simulated random testing of students, routine testing of teachers, testing of symptomatic students and teachers and the first quarantine policy where a full 14-day quarantine was imposed for a class in which any student or teacher received a positive test (RADT or PCR). Based on the mean across 100 simulation runs for 182 days each, with an estimated sensitivity (compared to PCR) of the RADT of 0.41 (IQR: 0.39 – 0.42) as compared to the true (i.e., modelled) sensitivity of 0.40 and an estimated specificity of POC test of 0.99 (IQR: 0.99 – 0.99) as compared to the true specificity of 0.98, we expected that the number of infections and tests would be sufficient in one single school to estimate the accuracy of RADT with acceptable precision. An additional school was added to support generalizability of the findings, as well as to allow exploration of the secondary objectives.
