## Supplementary material for "Evaluation of real-life use of Point-Of-Care Rapid Antigen TEsting for SARS-CoV-2 in schools (EPOCRATES)": Comparison of outbreaks to other high schools in Montreal using public health data during the same period

***Supplementary Appendix B: Comparison of outbreaks to other high schools in Montreal using public health data during the same period***

Data from Montréal Public Health showed that in other Montréal high schools (n=177), a range of 1 to 16 exposures and outbreaks per school (median: 1; IQR: 2) were observed during the study period, for a total of 358 outbreaks, one affecting two different schools. Schools declared that 1 to 52 cases (median: 4; IQR 5) were linked to an outbreak, for a total of 1181 cases. A range of 1 to 25 classes (median: 2, IQR: 3) were involved in outbreaks (n=161), for a total of 447 classes. Outbreaks at ESCL and PSNM comprised, on average, 3 and 2 cases, respectively. ESCL and PSNM had three outbreaks, with nine and seven students involved, from four and two groups, respectively, during the same period.

Despite active surveillance of SARS-CoV-2 infection through this study, there was no difference in outbreaks observed between participating schools and the rest of the Montréal high schools. However, outbreaks declared in other schools had a lower proportion of asymptomatic cases (31.8%) compared to ESCL (55.6%) and PSNM (85.7%). Participating schools had a lower proportion of cases linked to an outbreak present in school while contagious (28.6% and 6.7%), compared to the average in other schools of Montréal (n=241; 36.5%). Interestingly, data showed that 66.0% of cases linked to an outbreak in other high schools tested positive or started their symptoms within seven days of their first exposure (Supplementary Figure). Furthermore, 51.0% of the 741 cases linked to an outbreak who went to school while contagious were only processed by the Public Health team, due to capacity, after the recommended first testing date.

Using the number of classes where an outbreak occurred, assuming on average that one student is in class while contagious, we estimated that 50,010 high school students were isolated during the study period in other high schools which, with 14 days of isolation, leads to an estimated 700,140 days or ~1,918 years of cumulative isolation. A safe, accelerated return to school could have possibly saved an estimated 350,070 days or ~959 years of cumulative isolation (Supplementary Table).
