## Supplementary material for "Evaluation of real-life use of Point-Of-Care Rapid Antigen TEsting for SARS-CoV-2 in schools (EPOCRATES)": Predicted days of isolation

***Supplementary Table. Predicted days of isolation***

| **January 25 to June 10 2021** | **Average students per group** | **Groups** | **Total students put in isolation (instances)** | **Days of isolation (14 days)** | **Days of isolation (7 days)** |
| --- | --- | --- | --- | --- | --- |
| High school | 30 | 1667 | 50010 | 700140 | 350070 |
