## Supplementary material for "Evaluation of real-life use of Point-Of-Care Rapid Antigen TEsting for SARS-CoV-2 in schools (EPOCRATES)": Distribution of non-index cases linked to an outbreaks according to time to symptoms

Distribution of non-index cases linked to an outbreak according to the time (days) needed for their symptoms to appear or to test positive

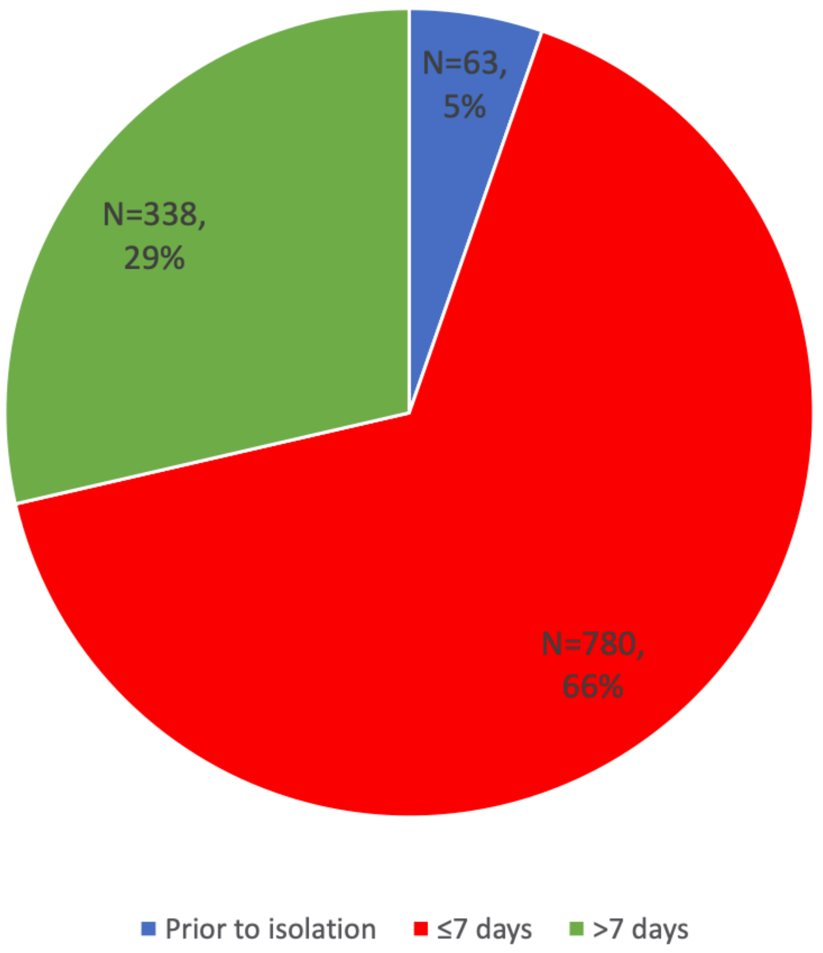
